# Introduction and sustained-transmission risk across DRC health zones during the Bundibugyo virus disease outbreak

**DOI:** 10.64898/2026.06.11.26355237

**Authors:** Reagan Luvande Okingo, Berthe Amélie Iroungou, Eugenio Valdano

## Abstract

During the ongoing Bundibugyo ebolavirus disease outbreak in the Democratic Republic of the Congo, we quantify introduction risk and sustained-transmission potential across the country. This identifies priority zones far from currently affected areas, where rapid amplification could follow introduction and response efforts should be focused.

## Body

An outbreak of Bundibugyo virus disease is ongoing in Ituri province, Democratic Republic of the Congo (DRC). Insecurity, high population mobility, limited healthcare capacity, and the absence of a vaccine pose challenges to response^1^. As of June 4, confirmed cases in Nord-Kivu and Sud-Kivu indicate that the spread has gone beyond the initial focus. Rapid risk assessment should therefore identify which spatial communities in DRC are most likely to receive cases and amplify the outbreak. This requires estimating introduction risk and, more importantly, the potential for sustained onward transmission, despite limited data on human mobility, contact patterns, disease transmissibility, tracing and isolation capacity.

We used the method developed in Ref.^2^ to estimate, for each DRC health zone (HZ), the potential for an introduced case to generate sustained transmission (hereafter, sustained-transmission potential). We used available population and mobility data to parameterize spatial transmission^3^, testing a broad range of assumptions on disease transmissibility and on how local and between-community contact patterns can be inferred from these data. We ranked HZs by sustained-transmission potential and compared them with introduction-risk ranks estimated from mobility data^4^. Unlike risk estimates, risk rankings were robust across scenarios and assumptions and can therefore inform action despite limited knowledge of transmissibility and contact patterns. Details in the Appendix.

Introduction risk was highest in HZs close to the reported foci, specifically in Ituri, Nord-Kivu, and Haut-Uele, consistent with expected mobility patterns (Fig. 1A,B). Non-negligible risk of introduction also appeared in distant urban centers, including the Kinshasa area. By contrast, high sustained-transmission potential was distributed across DRC, including Kinshasa, Kasaï-Oriental, Kongo-Central, Tshopo, Nord-Kivu, Haut-Katanga, Sud-Kivu, Kasaï, and Kasaï-Central (Fig. 1C,D). Notably, several are far from the current outbreak. Crossing the two rankings identified eight HZs that combined high introduction risk and high sustained-transmission potential (Fig. 1E, F): Binza Ozone and Binza Meteo in Kinshasa; Karisimbi in Nord-Kivu; Kadutu and Ibanda in Sud-Kivu; and Makiso-Kisangani, Kabondo, and Mangobo in Tshopo.

**Figure 1.**
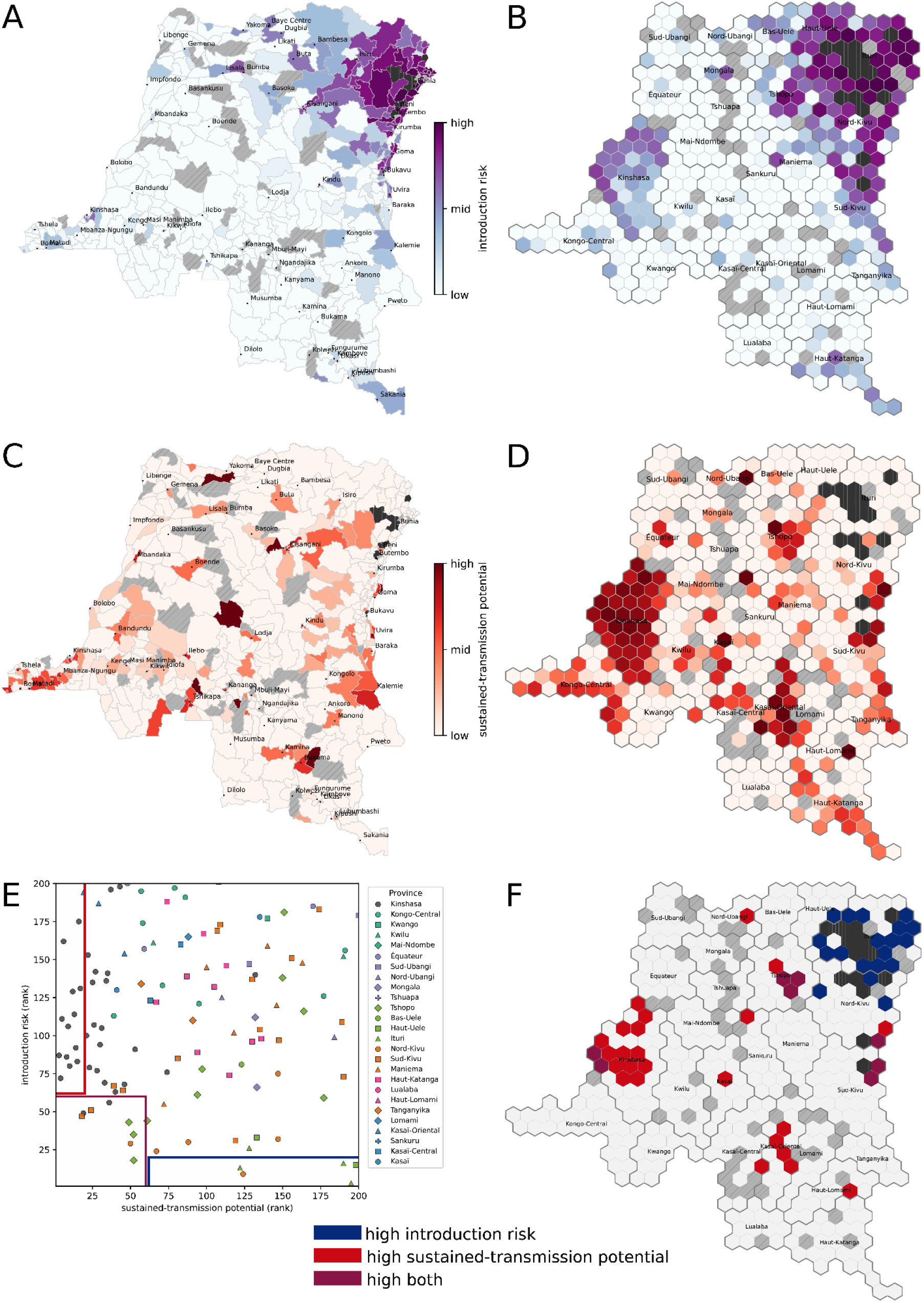
Introduction risk and sustained-transmission potential across health zones. A) Introduction to risk in each health zone. Health zones with reported confirmed cases are in dark gray. Health zones with no data are in light gray. B) Cartogram representation of A. C) Sustained-transmission potential in each health zone. D) Cartogram representation of D. E) Scatter plot of the ranked introduction risk and sustained-transmission potential for each health zone. Provinces are identified by dot color/shape. Low rank means high risk. Blue rectangle identifies health zones with high introduction risk but relatively low sustained-transmission potential. The red rectangle identifies high sustained-transmission potential but relatively low introduction risk. Purple rectangle identifies health zones with high introduction risk and high sustained-transmission potential. Population data are from worldpop.org. Mobility data are from flowminder.org. Methodology, additional data sources, additional results and tabled values are available in the Appendix.

These results separate importation from amplification risk: health zones near the affected area are more likely to receive cases, but not necessarily to sustain transmission. Our estimates therefore differ from Ref.^5^ by identifying areas of concern farther from the affected zone. This difference highlights the value of multiple, independent risk assessments in providing robust and outcome-specific guidance. Our results warrant preparedness in health zones with high sustained-transmission potential but lower introduction risk, particularly in Kinshasa and other urban centers, despite their distance from the outbreak. This is relevant because several high-amplification areas are in Kinshasa, Kasaï, and Tshopo, where preparedness may be more feasible than in parts of eastern DRC despite heterogeneous healthcare access, accessibility constraints, and low-to-moderate insecurity outside Kinshasa. Response planning should therefore not focus solely on areas closer to the outbreak, where the risk is dominated by introduction risk. Finally, health zones high on both dimensions (purple in Fig. 1F) should be prioritized for reinforced surveillance, rapid alert investigation, and pre-positioning of response capacity.

## Data Availability

All data produced are available online at https://data.humdata.org/ and https://www.worldpop.org/

