## Appendix for "Introduction and sustained-transmission risk across DRC health zones during the Bundibugyo virus disease outbreak"

### 1 Methods

#### 1.1 Parametrization of the reproduction operator

The reproduction operator $\mathbf{R}$ is defined in Ref. [1, 2]. Given $n$ distinct spatial communities (here the $n = 516$ health zones in the Democratic Republic of the Congo), $R_{ij}$ $(i, j = 1, \cdots n)$ is defined as the expected numbers of secondary infections that an infected resident of $j$ generates in $i$. From $\mathbf{R}$ it is possible to define the *reference reproduction ratio* $R^{\text{ref}}$ [1] and the community-specific outbreak reproduction ratio [2] (see also Sec. 1.4. We parametrized $\mathbf{R}$ it from spatial mobility patterns with a similar approach as what we did in Ref. [3]:

$$R_{ij} = \beta N_i \sum_k A_{ik} A_{jk} \frac{\eta_k}{\Omega_k}; \tag{1}$$

$$\Omega_i = \sum_j N_j A_{ji}. \tag{2}$$

$N_i$ is the resident population of $i$, $A_{ij}$ is the expected fraction of time that a resident of $i$ spends in $j$ (normalization $\sum_j A_{ij} = 1$), $\Omega_i$ is the expected number of people present in $i$ at a given time, $\eta_i$ is the transmissibility in community $i$, relative to an overall transmissibility level that is tuned by $\beta$ and fixed by the reproduction ratio.

##### Parametrization of A

We informed $N_i$ with population data from [4], aggregating high-resolution estimates to the level of health zones. Then $F_{ij}$ be the raw monthly relocation flux from $i$ to $j$, provided by Flowminder [5]. By convention, let us impose $F_{ii} = 0$. We defined the normalized relocation flux: $f_{ij} = F_{ij}/N_i$ and the total normalized outgoing flow: $f_i = \sum_j f_{ij}$. Then,

$$A_{ij}(\lambda) = \frac{(1 - f_i)\delta_{ij} + \lambda f_{ij}}{1 + (\lambda - 1) f_i}. \tag{3}$$

Note that the normalization is correct: $\sum_j A_{ij}(\lambda) = 1$. $\lambda$ is the translation factor from relocation fluxes to recurrent mobility fluxes. $\lambda = 1$ means that relocation flows are treated as the fraction of the origin population effectively present in the destination during the month. $\lambda < 1$ means that relocations overstate transmission-relevant exposure. Finally, $\lambda > 1$ means that relocations understate exposure because they miss shorter recurrent mobility or multiple contacts created by travel.

##### Parametrization of $\eta$

Ebola transmission rate may be higher in urbanized settings [6, 7]. Therefore we tested different dependence on the population density. Let $\delta_i$ be the population density in $i$, then $\eta_i(\theta) \propto \delta_i^\theta$ (the proportionality constant is irrelevant since $\beta$ absorbs it). $\theta = 0$ means that transmissibility is constant across communities. $\theta = 1$ means that it grows linearly with population density. $\theta > 1$ ($\theta < 1$) it means it grows superlinearly (sublinearly) with population density. To estimate population density in a health zone we averaged population density at the level of $100m \times 100m$ tiles, provided in Ref. [4], weighted by the population of that tile, over the whole health zone. This gave the average population density ($\delta_i$) experienced by the average resident of the health zone $i$, which accounts for a possibly heterogeneous distribution of the population across density levels. Its formula thus is

$$\delta_i = \frac{\sum_{p \in i} n_p^2}{\sum_{p \in i} n_p}, \tag{4}$$

where $p \in i$ means summing over all the $100m \times 100m$ covering $i$.

##### Scenarios

In the scenarios, we explored all possible combinations of these values ($2,100$ in total):

- $R_0$: 10 equally spaced values in $R_0 \in [1.25, 3]$;
- $\lambda$: 10 logarithmically spaced values in $\lambda \in [0.1, 10]$;
- $\theta \in \{1/2, 3/4, 1, 5/4, 3/2, 7/4, 2\}$.

To translate $R_0$ into $\beta$ we set $R_0$ to be equal to the local reproduction ratio $R_{ii}$ in *Bunia* health zone, currently the one with the highest number of cases.

### 1.2 Relocation and population data

We used relocation data from Ref. [5] averaged from May 2025 to April 2026. Relocation fluxes smaller than 15 were not reported and had to be imputed. To do it, first we fitted a power law to the raw occurrence of flux values in the interval $[15, 100)$ (see Fig. 1). The fit gave the following power law exponent: $\text{flux}^{-1.46}$, which, as the figure shows, fitted the data well. We therefore used it to extrapolate. we imputed censored values by sampling them proportionally to $\text{flux}^{-1.46}$ with $\text{flux} \in [1, 15)$.

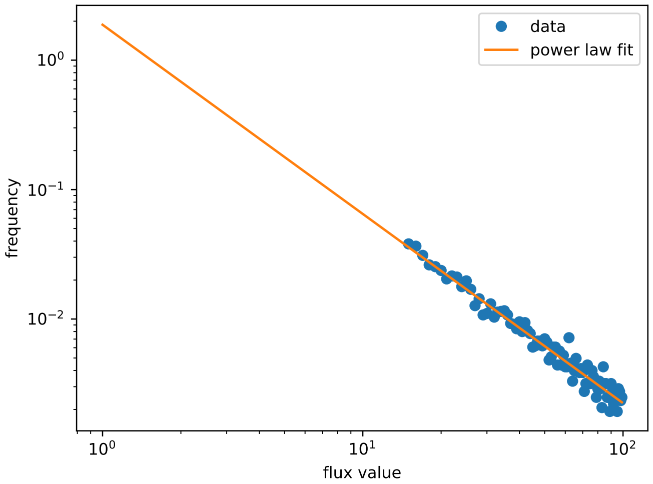


Figure 1: Power law fit of the raw occurrences of relocation flux data in the interval $[15, 100)$. The fit is $y = qx^m$ with $q = 1.90$ and $m = -1.46$.

### 1.3 Estimating relative introduction risk

Let $y_i$ be the cumulated incidence in HZ $i$, retreived from Ref. [8]. Let $\mathcal{S}$ be the set of possible sources, i.e., those for which $y_i > 0$. Let $F_i = \sum_{j \in \bar{\mathcal{S}}} F_{ij}$ the total flux from $i$ which does not go to sources (let $\bar{\mathcal{S}}$ be the complement of $\mathcal{S}$). Then, following Ref. [9], the relative risk of introduction to $i$ is

$$\rho_i = \frac{\sum_{j \in \mathcal{S}} F_{ji} y_j}{\sum_{j \in \mathcal{S}} F_j y_j}. \tag{5}$$

#### 1.4 Estimating sustained-transmission potential

Given a specific reproduction operator $\mathbf{R}$ we computed the outbreak reproduction ratio $R_i^{\mathrm{ob}}$ in each health zone using Ref. [2]. The outbreak reproduction ratio in community $i$ encodes the potential that an introduction of the pathogen in that community leads to a large-scale outbreak. As explained in the reference, the outbreak reproduction ratio can be estimated from the probability $p_i$ that a branching process seeded in $i$ does not go extinct: $R_i^{\mathrm{ob}} = -\log(1-p_i)/p_i$. In turn, the probability solves the equation $p_i = 1 - \exp(-\sum_j R_{ji} p_j)$.

Finally, we define the sustained-transmission potential in health zone $i$ as the median value of the rank of its value of $R_i^{\mathrm{ob}}$ across scenarios. In Sec. 2.1 we show that the rank value is stable across scenarios.

We also remark that the sustained-transmission potential in its ranked form can also be estimated directly from $p_i$ given that the rank measured on $R_i^{\mathrm{ob}}$ is the same as that measured on $p_i$ when $R_i^{\mathrm{ob}} \geq 1$.

### 2 Supplementary results

#### 2.1 Robustness of the rank estimates for the sustained-transmission potential

Figure 2 confirms that the ranked values of the sustained-transmission potential are stable across scenarios. Rank does not sensibly change across scenarios for the $\approx 120$ highest-risk HZ. Even for those with median risk between $\approx 120 - 200$, higher risk values are never smaller than $\approx 100$. See Tab. 1 for the exact values. This proves that our definition of sustained-transmission potential does not depend on the assumptions on the parameters defining spatial transmission.

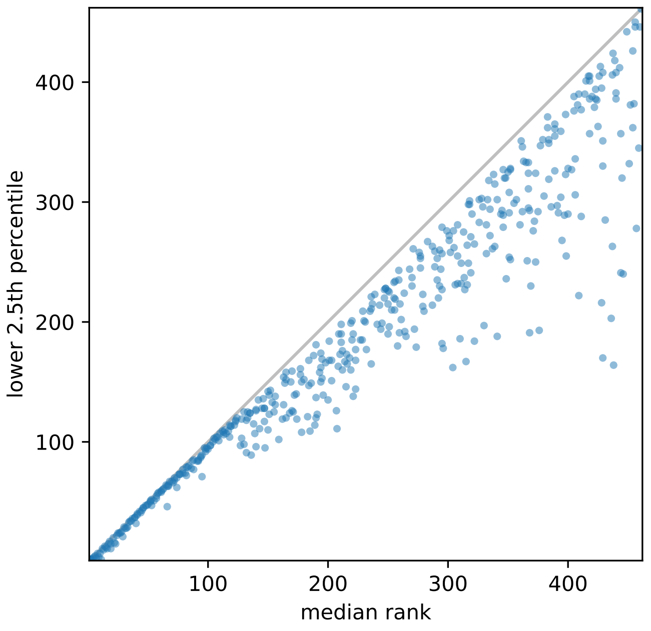


Figure 2: Robustness of the estimate of the sustained-transmission potential. Each dot represents a health zone. The $x$ axis is the estimated median rank of the sustained-transmission potential across scenarios (1 meaning highest value). The $y$ axis is the top 2.5th percentile of the rank.

#### 2.2 Reported risk values

Table 1 reports values of the rank of the sustained-transmission potential and the introduction risk as well as the categories displayed in Fig. 1 of the main text.

Table 1: Ranked sustained-transmission potential and introduction risk by health zone. HZ ranking $200th$ or higher in both median sustained-transmission potential (sust.-trans.) and introduction risk (introd.). Column cat. identifies HZ that are considered at high risk for sustained-transmission potential only (hs_li), introduction risk only (ls_hi) or both (hs_hi) – see Fig. 1 of the main text for the assignment.

| HZ (province) | sust.-trans. (median) | sust.-trans. (2.5th perc.) | introd. (rank) | cat |
|---|---|---|---|---|
| Bulu (Sud-Ubangi) | 200 | 135 | 179 | none |
| Watsa (Haut-Uele) | 198 | 166 | 15 | ls_hi |
| Tchomia (Ituri) | 195 | 169 | 3 | ls_hi |
| Gombe-Matadi (Kongo-Central) | 191 | 123 | 156 | none |
| Mandima (Ituri) | 190 | 181 | 16 | ls_hi |
| Kabare (Sud-Kivu) | 190 | 120 | 73 | none |
| Mosango (Kwilu) | 190 | 137 | 152 | none |
| Kaziba (Sud-Kivu) | 189 | 114 | 109 | none |
| Yakoma (Nord-Ubangi) | 184 | 147 | 99 | none |
| Bafwagbogbo (Tshopo) | 177 | 156 | 59 | none |
| Inga (Kongo-Central) | 177 | 161 | 126 | none |
| Nundu (Sud-Kivu) | 174 | 119 | 183 | none |
| Bolenge (Équateur) | 170 | 126 | 185 | none |
| Basoko (Tshopo) | 164 | 149 | 116 | none |
| Lubutu (Maniema) | 163 | 154 | 148 | none |
| Shabunda (Sud-Kivu) | 156 | 138 | 151 | none |
| Ubundu (Tshopo) | 151 | 139 | 181 | none |
| Isangi (Tshopo) | 150 | 142 | 138 | none |
| Nyantende (Sud-Kivu) | 148 | 95 | 97 | none |
| Kayna (Nord-Kivu) | 147 | 128 | 32 | none |
| Walikale (Nord-Kivu) | 147 | 117 | 75 | none |
| Punia (Maniema) | 146 | 136 | 120 | none |
| Saramabila (Maniema) | 140 | 126 | 159 | none |
| Kenge (Kwango) | 140 | 96 | 177 | none |
| Manika (Haut-Katanga) | 136 | 89 | 98 | none |
| Lemera (Sud-Kivu) | 135 | 124 | 104 | none |
| Mweka (Kasaï) | 135 | 124 | 178 | none |
| Isiro (Haut-Uele) | 133 | 120 | 33 | none |
| Yamaluka (Mongala) | 133 | 125 | 66 | none |
| Lisala (Mongala) | 132 | 118 | 112 | none |
| Maluku 1 (Kinshasa) | 132 | 91 | 140 | none |
| Mumbunda (Haut-Katanga) | 130 | 125 | 96 | none |
| Kimbi Lulenge (Sud-Kivu) | 130 | 98 | 137 | none |
| Nia Nia (Ituri) | 128 | 103 | 26 | none |
| Gemena (Sud-Ubangi) | 128 | 119 | 147 | none |
| Beni (Nord-Kivu) | 124 | 120 | 9 | ls_hi |
| Buta (Bas-Uele) | 123 | 118 | 81 | none |
| Mambasa (Ituri) | 122 | 114 | 13 | ls_hi |
| Katana (Sud-Kivu) | 119 | 113 | 31 | none |
| Kalima (Maniema) | 118 | 114 | 102 | none |
| Lubumbashi (Haut-Katanga) | 115 | 107 | 74 | none |
| Rwashi (Haut-Katanga) | 113 | 108 | 146 | none |
| Gbadolite (Nord-Ubangi) | 110 | 107 | 125 | none |
| Kamituga (Sud-Kivu) | 109 | 101 | 173 | none |
| Fizi (Sud-Kivu) | 107 | 104 | 169 | none |
| Kenya (Haut-Katanga) | 105 | 103 | 132 | none |
| Alunguli (Maniema) | 100 | 94 | 145 | none |
| Likasi (Haut-Katanga) | 98 | 92 | 167 | none |
| Bafwasende (Tshopo) | 97 | 95 | 78 | none |
| Kitona (Kongo-Central) | 95 | 71 | 121 | none |
| Lubunga (Tshopo) | 94 | 87 | 61 | none |
| Kampemba (Haut-Katanga) | 94 | 88 | 89 | none |

| HZ (province) | sust.-trans. (median) | sust.-trans. (2.5th perc.) | introd. (rank) | cat |
|---|---|---|---|---|
| Nyemba (Tanganyika) | 91 | 84 | 110 | none |
| Nyirangongo (Nord-Kivu) | 88 | 77 | 30 | none |
| Mwene-Ditu (Lomami) | 88 | 85 | 165 | none |
| Tshamilembe (Haut-Katanga) | 87 | 84 | 139 | none |
| Kimpese (Kongo-Central) | 86 | 78 | 191 | none |
| Lukonga (Kasaï-Central) | 83 | 79 | 160 | none |
| Ruzizi (Sud-Kivu) | 81 | 74 | 85 | none |
| Kizu (Kongo-Central) | 79 | 77 | 197 | none |
| Nsele (Kinshasa) | 74 | 62 | 76 | none |
| Katuba (Haut-Katanga) | 74 | 70 | 188 | none |
| Kindu (Maniema) | 72 | 70 | 55 | none |
| Nsona-Pangu (Kongo-Central) | 71 | 66 | 133 | none |
| Rutshuru (Nord-Kivu) | 67 | 64 | 24 | none |
| Kamalondo (Haut-Katanga) | 67 | 63 | 122 | none |
| Kikwit-Nord (Kwilu) | 65 | 64 | 161 | none |
| Katoka (Kasaï-Central) | 63 | 61 | 123 | none |
| Tshopo (Tshopo) | 61 | 58 | 44 | none |
| Mbandaka (Équateur) | 59 | 58 | 157 | none |
| Moanda (Kongo-Central) | 58 | 57 | 174 | none |
| Kalemie (Tanganyika) | 57 | 55 | 134 | none |
| Mbanza-Ngungu (Kongo-Central) | 57 | 53 | 195 | none |
| Mont Ngafula 1 (Kinshasa) | 53 | 47 | 91 | none |
| Makiso-Kisangani (Tshopo) | 52 | 51 | 18 | hs_hi |
| Kabondo (Tshopo) | 52 | 50 | 35 | hs_hi |
| Karisimbi (Nord-Kivu) | 50 | 48 | 29 | hs_hi |
| Mangobo (Tshopo) | 49 | 47 | 43 | hs_hi |
| Wangata (Équateur) | 48 | 47 | 200 | none |
| Mont Ngafula 2 (Kinshasa) | 46 | 45 | 68 | none |
| Nzaba (Kasaï-Oriental) | 46 | 45 | 154 | none |
| Bagira (Sud-Kivu) | 45 | 43 | 64 | none |
| Biyela (Kinshasa) | 43 | 42 | 198 | none |
| Kanzala (Kasaï) | 41 | 40 | 130 | none |
| Gombe (Kinshasa) | 40 | 32 | 63 | none |
| Uvira (Sud-Kivu) | 39 | 37 | 67 | none |
| Matadi (Kongo-Central) | 39 | 37 | 113 | none |
| Police (Kinshasa) | 37 | 36 | 196 | none |
| Binza Meteo (Kinshasa) | 36 | 34 | 56 | hs_hi |
| Kikimi (Kinshasa) | 35 | 34 | 141 | none |
| Kimbanseke (Kinshasa) | 34 | 33 | 136 | none |
| Limete (Kinshasa) | 33 | 29 | 69 | none |
| Selembao (Kinshasa) | 31 | 28 | 105 | none |
| Kingabwa (Kinshasa) | 30 | 29 | 94 | none |
| Bonzola (Kasaï-Oriental) | 29 | 21 | 187 | none |
| Kokolo (Kinshasa) | 28 | 25 | 77 | none |
| Kinsenso (Kinshasa) | 27 | 24 | 142 | none |
| Masina 1 (Kinshasa) | 26 | 24 | 107 | none |
| Masina 2 (Kinshasa) | 25 | 24 | 135 | none |
| Kadutu (Sud-Kivu) | 24 | 22 | 51 | hs_hi |
| Kasa-Vubu (Kinshasa) | 23 | 15 | 79 | none |
| Kalamu 2 (Kinshasa) | 22 | 16 | 153 | none |
| Lemba (Kinshasa) | 21 | 20 | 100 | none |
| Binza Ozone (Kinshasa) | 19 | 15 | 49 | hs_hi |
| Diulu (Kasaï-Oriental) | 19 | 11 | 194 | hs_li |
| Ibanda (Sud-Kivu) | 18 | 17 | 47 | hs_hi |
| Ndjili (Kinshasa) | 17 | 15 | 131 | hs_li |
| Makala (Kinshasa) | 16 | 11 | 175 | hs_li |
| Kalamu 1 (Kinshasa) | 15 | 12 | 83 | hs_li |
| Matete (Kinshasa) | 14 | 13 | 92 | hs_li |
| Barumbu (Kinshasa) | 13 | 10 | 114 | hs_li |
| Kingasani (Kinshasa) | 12 | 11 | 129 | hs_li |
| Kintambo (Kinshasa) | 10 | 7 | 80 | hs_li |

| HZ (province) | sust.-trans. (median) | sust.-trans. (2.5th perc.) | introd. (rank) | cat |
|---|---|---|---|---|
| Ngir Ngiri (Kinshasa) | 9 | 4 | 106 | hs_li |
| Lingwala (Kinshasa) | 8 | 7 | 86 | hs_li |
| Bumbu (Kinshasa) | 6 | 5 | 162 | hs_li |
| Ngaba (Kinshasa) | 5 | 4 | 111 | hs_li |
| Kinshasa (Kinshasa) | 4 | 3 | 72 | hs_li |
| Bandalungwa (Kinshasa) | 3 | 2 | 87 | hs_li |
